# Evaluation of *HOTAIR*, *HOXD8*, *HOXD9*, *HOXD11* Gene Expression Levels in Turkish Patients with Acute Myeloid Leukemia and Chronic Myeloid Leukemia: A Single Center Experience

**DOI:** 10.1101/2023.09.26.23296073

**Authors:** Esma Saraymen, Yakut Erdem, Hilal Akalın, Nazife Taşçıoğlu, Berkay Saraymen, Serhat Çelik, Yeşim Özdemir, Leylagül Kaynar, Mustafa Çetin, Yusuf Özkul

**Affiliations:** Central Research Facility Application and Research Center, Abdullah Gül University, Kayseri, Türkiye; Betül-Ziya Eren Genome and Stem Cell Center, Erciyes University, Kayseri, Türkiye; Department of Medical Genetics, Faculty of Medicine, Erciyes University, Kayseri, Türkiye; ERNAM-Nanotechnology Research and Application Center, Erciyes University, Kayseri, Türkiye; Yenimahalle Training and Research Hospital, Yıldırı Beyazıt University, Ankara, Türkiye; Department of Medical Genetics, Faculty of Medicine, Üsküdar University, İstanbul, Türkiye; Department of Hematology, Faculty of Medicine, İstanbul Medipol University, İstanbul, Türkiye; Department of Hematology, Medstar Antalya Hospital, Antalya, Türkiye

**Keywords:** *HOTAIR*, *HOXD8*, *HOXD9*, *HOXD11*, AML, CML

## Abstract

**BACKGRAUND:** Homeobox (HOX) transcript antisense RNA (HOTAIR) and *HOX* genes are reported to be more expressed in various cancers in humans in recent studies. The role of *HOTAIR* and *HOXD* genes in acute myeloid leukemia (AML) and chronic myeloid leukemia (CML) is not well known.

**MATERIALS AND METHODS:** In this study, expression levels of *HOXD8*, *HOXD9* and *HOXD11* from *HOXD* gene family and *HOTAIR* were determined from peripheral blood samples of 30 AML and 30 CML patients and 20 healthy volunteers by quantitative Real Time PCR.

**RESULTS:** We determined that the expression levels of *HOXD9* and *HOXD11* in the AML patients were significantly lower than the control group (p<0.001 and p=0.002, respectively). There was no significant difference in the expression levels of *HOTAIR* and *HOXD8* when compared to the control group. In the CML patients there was a significant increase in the expression level of *HOTAIR* when compared to the control group (p=0.002). The expression levels of *HOXD9* and *HOXD11* were found to be significantly lower than the control group (p<0.001).

**CONCLUSION:** Our study showed that *HOTAIR* may not be a biomarker in the diagnosis and is not significantly correlated with the clinicopathological prognostic characteristics of AML. Additionally; it can be said that *HOTAIR* is oncogenic by suppressing the expression of *HOXD9* and *HOXD11* but not *HOXD8* in CML patients. The expression profiles of *HOTAIR* may be a potential biomarker in the diagnosis of CML patients in predicting and monitoring drug resistance.

## Introduction

Acute myeloid leukemia (AML) is a heterogeneous clonal disorder of hematopoietic stem cells that lose normal differentiation ability. The disease is characterized by various cytogenetic and molecular abnormalities with different prognoses and gene expression profiles. It is the most common type of leukemia in adults and has the lowest survival rate among all leukemias. The prevalence of AML is 3.8 per 100000 people, and increases to 17.9 per 100000 adults aged 65 years and over. The most appropriate treatment should be chosen according to the patient’s genetic profile. For this purpose, long non-coding (lnc) RNAs have recently been used for biomarkers in the diagnosis of cancer (1).

Chronic myeloid leukemia (CML) is a myeloproliferative disorder of hematopoietic stem cells carrying the Philadelphia chromosome and oncogenic *BCR-ABL1* fusion gene. The peptide p210, the product of the *BCR-ABL*, has structurally activated tyrosine kinase activity that is involved in the pathogenesis of CML (2). CML is a malignancy that occurs in 1 or 2 cases in approximately 100000 people in adults every year and it is mostly seen between 25-60 years of age (3). Studies have been conducted to investigate the profile of micro RNA and lnc RNA sequences that have been validated at different stages of CML to better understand the disease and to improve therapeutic intervention in CML patients (4).

Lnc RNAs with a nucleotide number of more than two hundred are also referred to as macro RNA and long intergenic non-coding RNA and are not involved in protein generation (4). While many of the lnc RNAs have been reported to vary in expression levels in different cancers, there is insufficient information about their role in cancer development. Therefore, the explanation of the mechanisms of these molecules is one of the current research topics. Although the function of lnc RNAs is not fully known, increasing numbers of lnc RNA and accumulated evidence for its involvement in many biological processes have shown that they have important functions in normal and malignant cells (5). For this purpose, lnc RNAs have recently been used for biomarkers in the diagnosis of cancer (6). It has been stated that the expression profiles of lnc RNAs are associated with recurrent mutations, clinical features and outcomes in AML, some of these lnc RNAs may have a functional role in leukemogenesis and may also be used as biomarkers in AML (7). Studies have been conducted to investigate the profile of micro RNA and lnc RNAs sequences that have been validated at different stages of CML to better understand the disease and to improve therapeutic intervention in CML patients (8).

Homeobox (HOX) transcript antisense RNA (HOTAIR) was first introduced in 2007 by Rinn et al. as a lnc RNA which suppresses expression of *HOXD* genes on the 2nd chromosome. (6). The relationship between lncRNAs and polycomb repressive complex 2 (PRC2), including *HOTAIR*, is considered a common mechanism in the gene silencing event in epigenetic regulation. *HOTAIR* binding is required to direct PRC2 to specific regions of the genome. Thus, PRC2 correlates gene expression and silences epigenetically (9). It is clear that *HOTAIR* reprogramming chromatin to promote metastasis and investigates the molecular mechanisms of tumor formation, metastasis and drug resistance (10).

Because *HOX* genes play a critical regulatory role in many cellular processes, changes in the expression of these genes play an important role in the development of cancer by affecting many mechanisms such as proliferation, growth and differentiation in mutations (11). Studies have reported changes in the expression of HOX genes in tumors such as lung carcinoma, neuroblastoma, glioma, ovarian cancer and leukemia (12).

HOXD gene contains between 9-11 genes arranged in a homologous sequence organization located on chromosome 2 (2q31) (13).

*HOXD8* is a gene belonging to the *HOX* gene family consisting of 2 exons, located on the 2nd chromosome (2q31.1). Deletions occurring in the entire *HOXD* gene group or at the 5’ end of this gene have been associated with severe limb and genital abnormalities. Negative and positive regulation of transcription from the RNA polymerase II promoter in the embryo plays a role in skeletal development processes. In addition to its regulatory effects during embryogenesis, it has been reported that this particular gene may play a role in adult urogenital system function (14).

*HOXD9* is one of the *HOXD* genes, which is part of the developmental regulatory system located in chromosome regions 2q31-2q37 (15). *HOXD9* is important for embryonic segmentation and limb development throughout development. This gene participates in the development and modeling of the forelimb and axial skeleton (16).

*HOXD11* is one of the belonging to the *HOX* gene family, which regulates the development and control of many cellular processes such as cell development and shaping, proliferation, migration, and apoptosis. It is a homeodomain consisting of 2 exons that encode a protein of 338 amino acids located on the 2nd chromosome (2q31.1) (17). It has been reported that the *HOXD11* plays a role in anterior dorsal morphology in mice (18).

There is no information in the literature about how *HOXD8*, *HOXD9* and *HOXD11* gene expression levels change in AML and CML. Therefore, we aimed to investigate the changes in the expression levels of these genes simultaneously with HOTAIR in relevant patient groups.

## Materials and Methods

### RNA extraction and quantitative real time polymerase chain reaction

Total RNA was isolated from four-mililiter EDTA blood samples using the Trizol method (19). cDNA synthesis was made with the transcriptor High Fidelity cDNA Synthesis Kit and PreAmp cDNA synthesis was performed using the Real Time ready cDNA Pre-Amp Master kit according to the manufacturer’s instruction (Roche Diagnostics). *Beta actin* as housekeeping gene was used for normalization in gene expression analysis. Sequences of specific primers belonging to *HOTAIR*, *HOXD8*, *HOXD9*, *HOXD11* and *Beta actin* genes are shown in Table 1. Gene expression measurements using the LightCycler 480 Probes Master kit was performed by LightCycler 480 II (Roche Diagnostics Ltd. Rotkreuz, Switzerland) polymerase chain reaction instrument. Analyzes were evaluated with LightCycler 480 Software (release 1.5.0 SP4). Each of the patient and control samples were repeated three times. Two negative controls and calibrators were used in each study.

**Table 1.**
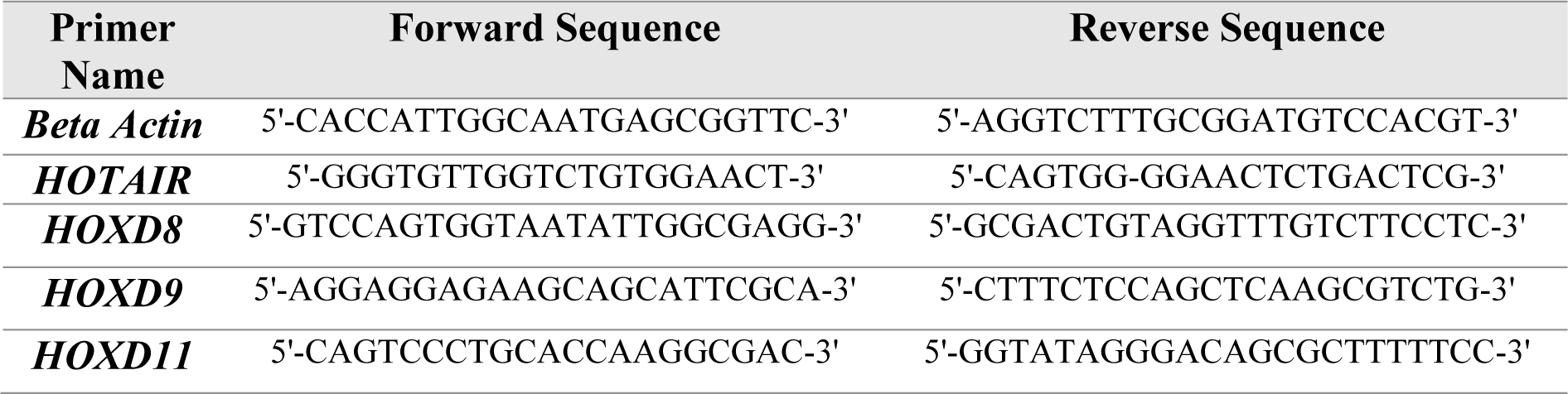
Sequences of specific primers belonging to *HOTAIR*, *HOXD8*, *HOXD9*, *HOXD11* and *Beta actin* genes.

The crossing points (Cp) values in the target genes of each sample were normalized to a ratio of the *Beta actin* housekeeping gene Cp of the same samples. In this manner, relative quantitation values were obtained (20).

### Statistical analysis

In our study, the statistical difference between the gene expression levels measured in patient and control samples was compared by using T test method and p values were calculated. The data were analyzed with SPSS 22.0 statistical program (Chicago, ILL, USA). p<0.05 was considered significant. Data are expressed as Mean ± Standard Error Mean (SEM).

## Results

### Clinical and demographic characteristics of patients

Newly diagnosed AML and CML patients, aged 18 to 75 years, who came to Erciyes University Medical Faculty Gevher Nesibe Hospital Medical Genetics Laboratory, consisted of 60 patients (30 AML, 30 CML), 35 of whom were male (20 AML, 15 CML) and 25 were female (10 AML, 15 CML), with a mean age of 42,07 ± 18,18 (AML) and 48.03 ± 19.90 (CML). 20 healthy individuals as a control group consisted of 13 of whom were male and 7 were female, with a mean age of 34,95 ± 9,59. Healthy volunteers who did not use any drug were choosen as controls. The age and sex distribution of the AML and CML patient groups were similar to the control group.

Classification of AML patients according to their clinicopathological characteristics are shown in Table 2 and clinical information of the CML patients are shown in Table 3.

**Table 2.**
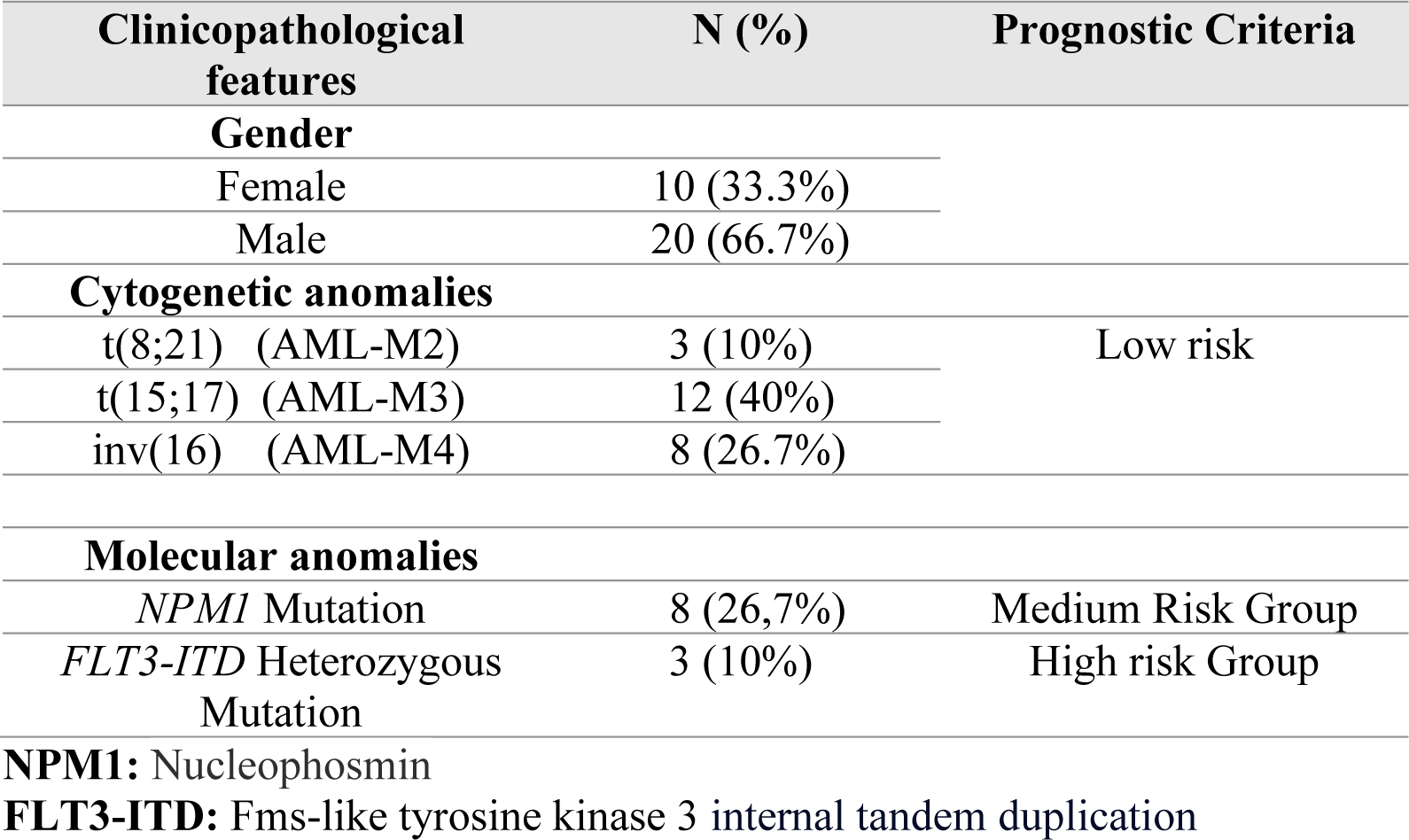
Clinicopathological characteristics of AML patients.

**Table 3.** The clinical information of CML patients.

| Patient | Gender | Medicine | Major<br>Molecular<br>Response | Prognostic Score |  |  |
| --- | --- | --- | --- | --- | --- | --- |
|  |  |  |  | EUTOS | SOKAL | HASFORD |
| P1 | Male | Imatinib | No | Low | Intermediate | Intermediate |
| P2 | Female | Imatinib | No | Low | Intermediate | Low |
| P3 | Male | Imatinib | No | Low | Low | Intermediate |
|  |  | Nilotinib | Yes |  |  |  |
| P4 | Male | Imatinib | Yes | Low | Intermediate | Low |
| P5 | Male | Imatinib | Yes | Low | High | Intermediate |
| P6 | Female | Imatinib | No | Low | High | Intermediate |
| P7 | Female | Imatinib | Yes | Low | Intermediate | Low |
| P8 | Female | Imatinib | No | Low | Low | Intermediate |
|  |  | Nilotinib | Yes |  |  |  |
| P9 | Female | Imatinib | No | Low | High | Intermediate |
| P10 | Female | Imatinib | Yes | Low | Low | Low |
| P11 | Male | Imatinib | No | Low | High | High |
| P12 | Female | Imatinib | Yes | Low | Intermediate | Low |
| P13 | Male | Imatinib | Yes | Low | Intermediate | Low |
| P14 | Female | Imatinib | Yes | Low | Intermediate | Intermediate |
| P15 | Male | Imatinib | Yes | Low | Low | Low |
| P16 | Female | Imatinib | No | Low | High | High |
| P17 | Male | Imatinib | Yes | Low | Intermediate | Intermediate |
| P18 | Female | Imatinib | Yes | Low | Low | Low |
| P19 | Female | Imatinib | No | Low | High | Intermediate |
| P20 | Male | Imatinib | No | Low | High | Intermediate |
| P21 | Female | Imatinib | Yes | Low | High | Low |
| P22 | Male | Imatinib | No | Low | Intermediate | Intermediate |
| P23 | Female | Imatinib | No | Low | Intermediate | Intermediate |
|  |  | Nilotinib | Yes |  |  |  |
| P24 | Male | Imatinib | Yes | Low | Intermediate | Low |
| P25 | Male | Imatinib | No | Low | High | Intermediate |
|  |  | Nilotinib | No |  |  |  |
| P26 | Male | Imatinib | No | Low | Intermediate | Intermediate |
| P27 | Female | Imatinib | Yes | Low | Intermediate | Intermediate |
| P28 | Male | Imatinib | No | Low | Low | Intermediate |
|  |  | Nilotinib | Yes |  |  |  |
| P29 | Male | Imatinib | Yes | Low | Intermediate | Intermediate |
| P30 | Female | Imatinib | No | Low | Low | Intermediate |
|  |  | Nilotinib | Yes |  |  |  |

**EUTOS:** European Treatment and Outcome Study for CML

### Comparison of target genes expression levels in AML, CML and control groups

The expression levels of *HOTAIR*, *HOXD8*, *HOXD9* and *HOXD11* were compared among AML patients, CML patients and healthy volunteers. The results were demonstrated in Figure 1.

**Figure 1.**
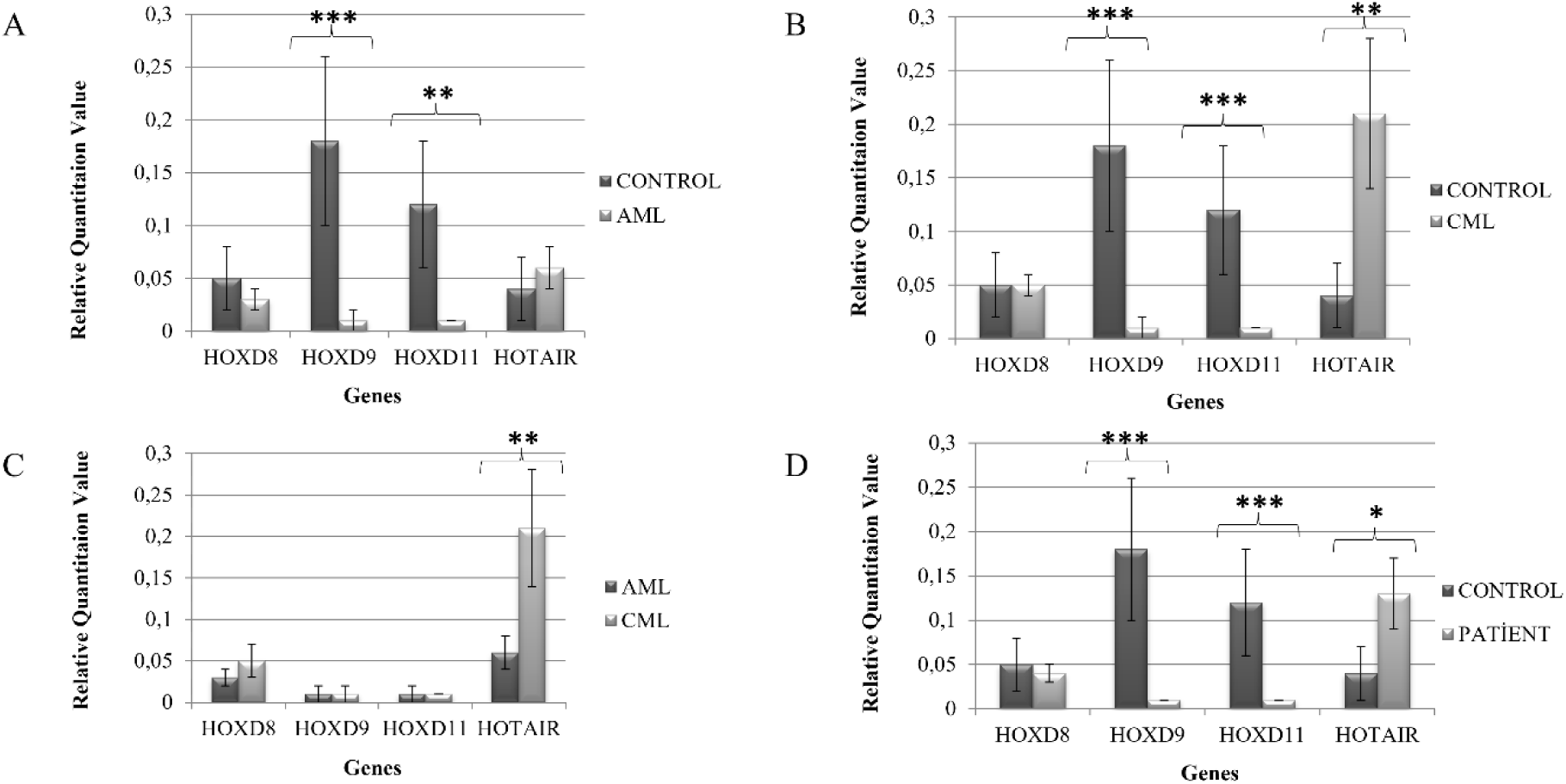
Comparison of *HOTAIR*, *HOXD8*, *HOXD9* and *HOXD11* expression levels in control AML, CML and patient (AML-CML) groups: *HOTAIR* expression levels were 0.04 ± 0.03, 0.06 ± 0.02, 0.21 ± 0.07 and 0.13 ± 0.04, respectively. *HOXD8* expression levels were 0.05 ± 0.03, 0.03 ± 0.01, 0.05 ± 0.02 and 0.04 ± 0.01, respectively. *HOXD9* expression levels were 0.18 ± 0.08, 0.01 ± 0.01, 0.01 ± 0.01 and 0.01 ± 0.00, respectively. *HOXD11* expression levels were 0.12 ± 0.06, 0.01 ± 0.01, 0.01 ± 0.00 and 0.01 ± 0.00, respectively. (Data shown as mean ± SEM. * p < 0.05, ** p < 0.01, *** p < 0.001).

When AML patients were compared with the control group, there was no statistically significant difference in the expression levels of *HOTAIR* and *HOXD8* (p=0.661 and p=0.242, respectively). The expression levels of *HOXD9* and *HOXD11* were significantly lower than the control group (p<0.001 and p=0.002, respectively) (Figure 1A).

In CML patients, the expression level of *HOTAIR* was found to be significantly higher than the control group (p=0.002). The expression levels of *HOXD9* and *HOXD11* were significantly lower than the control group (p<0.001). When the expression level of *HOXD8* was compared with the control group, there was no statistically significant difference (p=0.463) (Figure 1B).

*HOTAIR* expression level of CML patients was statistically higher than AML patients (p<0.01). There was no statistically significant difference in the expression levels of *HOXD8*, *HOXD9* and *HOXD11* between the both groups (p=0.396, p=0.725 and p=0.940, respectively). (Figure 1C).

*HOTAIR* expression level of the patient group (AML-CML) was statistically higher than the control group (p=0.032). The expression levels of *HOXD9* and *HOXD11* were significantly lower in the patient group than the control group (p<0.001). There was no statistically significant difference between the groups in the expression level of *HOXD8* (p=0.216) (Figure 1D).

## Discussion

While lnc RNAs contribute to epigenetic gene regulation, metastases and prognosis of solid tumors, its relationship with hematopoietic cancers has also started to be demonstrated in recent years (21). *HOTAIR*, characterized by increased expression of various human solid tumors, is a lnc RNA that is considered a potential cancer biomarker. Recent studies in many cancer types have shown that *HOTAIR* has a high expression level. *HOTAIR* has been reported to be overexpressed in various cancers such as colon, colorectal, gastric adenocarcinoma tissues, nasopharyngeal carcinoma, primary hepatocellular carcinoma, renal carcinoma cells in humans (22-24). *HOTAIR* suppresses the expression of *HOXD8*, *HOXD9*, *HOXD11* located in the *HOXD* locus of 2nd chromosome (6, 9).

There is insufficient information about the role of *HOTAIR* and *HOXD* in hematological malignancies. In addition, it is the first time study investigating the mRNA expression levels of *HOXD* genes such as *HOXD8*, *HOXD9* and *HOXD11* in patients with acute and chronic myeloid leukemia.

In studies conducted to investigate *HOTAIR* expression profile in AML patients and to evaluate their clinical significance; *HOTAIR* has been suggested that bad prognosis can be represented as a biomarker and may be a potential therapeutic target for the treatment of AML. The results showed that elevated *HOTAIR* expression level was significantly correlated with the high risk group of the disease (25-27).

Sayad et al. (28) reported that *HOTAIR* is not a biomarker for Iranian AML patients. They have suggested that *HOTAIR* expression level cannot be considered a definitive diagnostic and therapeutic biomarker for AML and should be confirmed by future studies and the correlation between *HOTAIR* and AML should still be investigated.

In our study, peripheral blood samples of 30 patients with AML and 20 healthy controls were evaluated. According to the findings, the expression levels of *HOTAIR* in AML patients were not statistically different when compared to the control group. Our investigations with AML patients is similar to the findings of Sayad et al. (28) and supports that *HOTAIR* expression level cannot be considered as a definitive diagnostic or therapeutic biomarker for AML.

A hybrid *BCR-ABL* oncogene is formed on chromosome 22, and its product, the p210 peptide, has constitutively activated tyrosine kinase activity that plays a role in the pathogenesis of CML (2). The widely used and highly effective treatment for CML is administration of imatinib mesylate, one of the tyrosine kinase inhibitors that inhibit the kinase activity of the *BCR-ABL* oncoprotein. Tyrosine kinase inhibitors targeting the *BCR-ABL* are the first-line therapy for most CML patients and have greatly improved the CML prognosis. However, imatinib resistance is emerging as a major challenge in the treatment of CML. Several molecular pathways have been associated with resistance to tyrosine kinase inhibitor therapy, but the precise mechanism for the development of drug resistance remains unclear. Therefore, in recent years, emphasis has been placed on the expression profiles of lnc RNAs, which can predict clinical outcomes in diagnosis, help prognostic classification of patients, predict and manage drug resistance (29).

In our study, it was seen that the expression levels of *HOTAIR* increased compared to the control group in the CML patients (p=0.002) In addition, clinical route and drug use of CML patients were followed. No significant difference was observed when the expression levels of the target genes measured at the time of diagnosis of CML patients were compared between patients who responded to treatment and those who did not and were resistant to imatinib.

Aghamohammadhossein et al. (30) have shown that *HOTAIR* expression levels in patients with positive CML for *BCR-ABL* increased compared to healthy individuals. They stated that changes in the expression levels of the *HOTAIR* may play a role in the biology of CML. In addition to this findings, in the present study, it was shown that *HOTAIR* expression levels increased while *HOXD9* and *HOXD11* expression levels decreased in CML patients compared to the healthy control group.

In previous studies, it was stated that *HOXD8*, *HOXD9*, *HOXD11* expression level profiles change in various cancer types (17, 31-35). In our study, for the first time, the expression levels of these genes were evaluated together with *HOTAIR* and demonstrated in AML and CML patients.

In the present study, it has been shown that *HOTAIR* epigenetically suppresses *HOXD9* and *HOXD11*, but *HOXD8* does not and may have an oncogenic effect in the development of CML.

## Conclusion

As a result, our study showed that *HOTAIR* may not be a biomarker in the diagnosis and is not significantly correlated with the clinicopathological prognostic characteristics of AML. Additionally; it can be said that *HOTAIR* is oncogenic by suppressing the expression of *HOXD9* and *HOXD11* but not *HOXD8* in CML patients. The expression profiles of *HOTAIR* may be a potential biomarker in the diagnosis of CML patients in predicting and monitoring drug resistance.

## Data Availability

All data produced in the present work are contained in the manuscript

## Authors’ contributions

**Esma Saraymen:** Data collection and processing, analysis and interpretation, literature search, critical reviews, writing; **Yakut Erdem:** data collection and processing, analysis and interpretation, literature search, critical reviews; **Hilal Akalım:** data collection and processing, analysis and interpretation; **Nazife Taşçıoğlu:** data collection and processing, analysis and interpretation; **Berkay Saraymen:** data collection and processing, literature search; **Serhat Çelik.** materials; data collection and processing; **Yeşim Özdemir:** data collection and processing; **Leylagül Kaynar:** supervision, materials, analysis and interpretation; **Mustafa Çetin:** supervision, materials, analysis and interpretation; **Yusuf Özkul:** concept, design, supervision, resource, materials, data collection and processing, analysis and interpretation, literature search, critical reviews.

## Availability of data

All data generated or analyzed during this study are included in the manuscript.

## Ethical Approval

Ethics Committee of Erciyes University Faculty of Medicine gave ethical approval for this work. (Approval number: 2015/222, Decision date: 08/05/2015).

## Consent of participants

Clinical and laboratory data and consent forms of the patients were also obtained from Erciyes University Medical Faculty Gevher Nesibe Hospital Medical Genetics Laboratory, The study was carried out in accordance with the principles of the Helsinki Convention on Human Rights.

